# Maternal genetic variants in kinesin motor domains prematurely increase egg aneuploidy

**DOI:** 10.1101/2024.07.04.24309950

**Authors:** Leelabati Biswas, Katarzyna M. Tyc, Mansour Aboelenain, Siqi Sun, Iva Dundović, Kruno Vukušić, Jason Liu, Vanessa Guo, Min Xu, Richard T. Scott, Xin Tao, Iva M. Tolić, Jinchuan Xing, Karen Schindler

## Abstract

The female reproductive lifespan depends on egg quality, particularly euploidy. Mistakes in meiosis leading to egg aneuploidy are common, but the genetic landscape causing this is not well understood due to limited phenotypic data. We identify genetic determinants of reproductive aging via egg aneuploidy using a biobank of maternal exomes linked with maternal age and embryonic aneuploidy data. We found 404 genes with variants enriched in individuals with high egg aneuploidy rates and implicate kinesin protein family genes in aneuploidy risk. Experimental perturbations showed that motor domain variants in these genes increase aneuploidy in mouse oocytes. A knock-in mouse model validated that a specific variant in kinesin *KIF18A* accelerates reproductive aging and diminishes fertility. These findings suggest potential non-invasive biomarkers for egg quality, aiding personalized fertility medicine.

**One sentence summary:** The study identifies novel genetic determinants of reproductive aging linked to egg aneuploidy by analyzing maternal exomes and demonstrates that variants in kinesin genes, specifically *KIF18A*, contribute to increased aneuploidy and accelerated reproductive aging, offering potential for personalized fertility medicine.

## Main text

Female fertility lies at the interface of multiple, complex organ systems and interwoven hormonal axes, all of which are governed by thousands of genes. At the convergence of this interplay is the egg. A female must have eggs of sufficient quantity and quality (*i.e.*, biological functionality) to successfully reproduce. One aspect of egg quality, known as egg euploidy—the presence of a correct chromosome complement— is a *sine qua non* for fertility and reproduction. Euploid eggs, once fertilized, can give rise to developmentally competent embryos; while aneuploid eggs typically cause a failure to implant or early miscarriage (*1–3*).

Currently, maternal age is the only clinical predictive biomarker for egg euploidy. Aging eggs lose chromosomal cohesion proteins that protect egg euploidy (*4–7*). As a result, age-dependent increases in egg aneuploidy taper the human female reproductive lifespan beginning in the early thirties and culminating to a near total loss of reproductive potential by the mid-forties, on average, prior to the normal onset of menopause (*8, 9*). However, there is considerable interindividual heterogeneity in female fertility and female reproductive lifespans (*10*). Recent studies examining age at natural menopause and female fertility further underscore that maternal genetic variants may contribute to this heterogeneity (*11*).

Elevated egg aneuploidy (EEA), a form of accelerated reproductive aging in which individuals have higher egg aneuploidy than expected for their maternal age, is a particular example of a pathological phenotypic variation affecting female fertility (*12*). The genetic basis of EEA is poorly understood, and the identification of causal genetic variants for EEA has been challenging for several reasons. First, egg aneuploidy phenotype data linked with maternal genetic sequencing information are limited. Assessment of an individual’s egg aneuploidy status can only occur in a fertility clinic, where the primary focus is on assisting with reproduction rather than investigating underlying causes. Consequently, patients undergoing this protocol do not routinely receive concurrent high-resolution genome or exome sequencing, which explains the rarity of linked genetic-phenotype data. Second, egg aneuploidy is a complex trait that arises from both normal age-dependent changes in eggs and genetic mechanisms. As a result, biologically validating specific genetic variants requires complex, multi-pronged model systems that combine multiple factors, such as advanced age and genetic variation. Therefore, the elucidation of the maternal genetic determinants of egg aneuploidy remains scarce to date.

Here, we overcome these limitations through a comprehensive multi-disciplinary approach. Using a bioinformatic strategy, we identify genes associated with EEA. By extending to experimental approaches in mice, we unravel novel roles of molecular motor proteins called kinesins in maintaining egg euploidy and shaping reproductive aging. Kinesins are a superfamily of 14 classes of oligomeric motor proteins that bind microtubules, usually have plus-end directed movement, and share a common motor domain (*13–15*). We report human variants in kinesin genes *KIF20A* and *KIF18A* associated with pathological EEA. KIF20A is a dimeric kinesin that transports the chromosomal passenger complex (CPC) in mitosis and meiosis, a function important for faithful chromosome segregation (*16–21*). KIF18A regulates microtubule dynamics and aneuploid cells are uniquely sensitive to its loss (*22–24*). We experimentally examined variants from human patients in mouse oocytes and show they perturb meiosis and elevate meiotic aneuploidy. Our findings not only offer insight into the basis of heterogeneity in reproductive aging but also provide opportunities for leveraging these mechanisms to optimize reproductive health across the female lifespan.

## Results

### Patient description and data quality

We previously collected de-identified maternal age and embryonic aneuploidy data from 753 White, non-Hispanic patients treated at Reproductive Medicine Associates of New Jersey. Using a robust nonlinear regression analysis, we selected 182 patients who had either high (43% to 100%; average 67%) or low (0% to 50%; average 11.1%) aneuploidy rate relative to their maternal ages. Whole-exome sequencing was performed on the 182 patients, followed by quality control, resulting in 178 patients with extreme aneuploidy rates (93 high, 85 low) for association studies, as previously described (*25*) (Fig. 1A, Table S1). The patients’ ages ranged from 24 to 43 years with a median of 37 years. The low-aneuploidy group had a median age of 37 years (IQR: 34-39) and the high-aneuploidy group had a median age of 36 years (IQR: 33-39). Here, we analyzed 494,448 variant sites, including 441,457 single nucleotide variants (SNVs) and 52,991 insertions/deletions (INDELs) (Supplemental Data S1).

**Figure 1.**
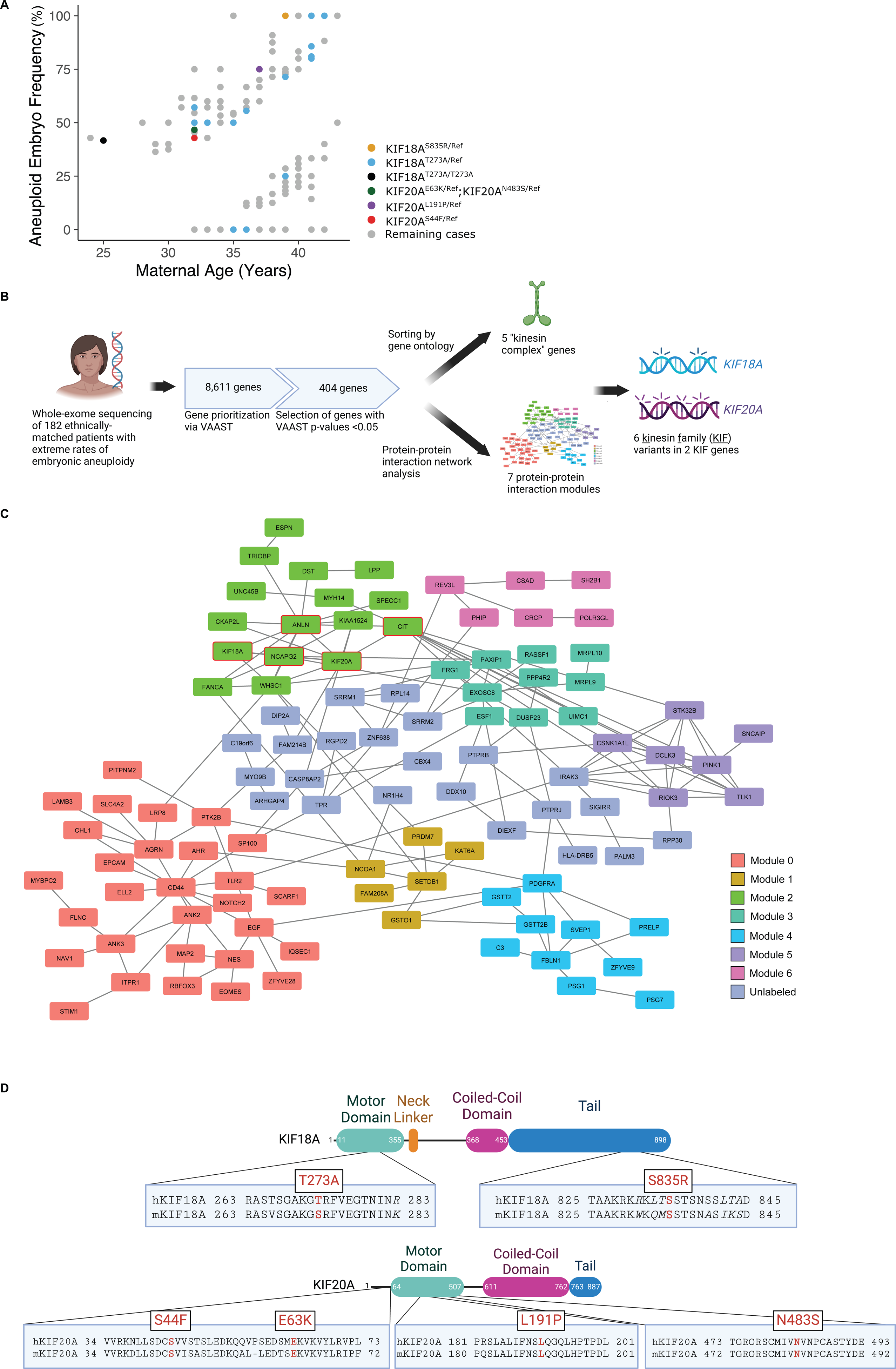
Bioinformatic approach to identify candidate aneuploidy genes. (A) Patient cohort and kinesin variant distribution. Colored dots indicate a genotype encoding the protein variants noted. “Ref” = reference allele. n = 182 patients. (B) Bioinformatic prioritization pipeline. Patients were divided into low-aneuploidy and high-aneuploidy cohorts and subjected to exome sequencing. Variants were identified and used for association analysis with VAAST. Genes identified by VAAST were used for gene ontology (GO) enrichment and protein–protein interaction network analyses. Genes that were enriched in the GO term “kinesin complex” and parts of a functional module were selected for functional studies in mouse oocytes. (C) Protein–protein interaction network of candidate genes. Genes are color-coded by their module assignments by PAPER. In module 2, genes belong to the “mitotic cell cycle process” GO term are circled in red. (D) Mutation diagram illustrating the protein consequences of EEA-enriched mutations in KIF18A and KIF20A. Local alignments of human KIF18A (hKIF18A) and mouse KIF18A (mKIF18A). Italicized residues are not conserved between human and mouse. Red, bolded residues are those substituted by an alternative allele in the high-aneuploidy individuals.

### VAAST and gene ontology analyses implicate kinesins in EEA

First, we used the program VAAST (*26*) to identify potentially causal variants in EEA patients. A total of 8,611 genes had positive scores, but no single gene achieved a genome-wide significance level. To assess the relevance of the genes that were prioritized by VAAST, we manually reviewed the top 50 genes for known mouse phenotypes reported in MouseMine (*27*). Out of the 50 genes, 27 had reported phenotypes in MouseMine and phenotypes for 13 of those genes were highly relevant in the context of this study (infertility, embryonic defects, prenatal lethality, *etc*.) (Table S2). We did not further pursue those 13 genes because we sought to focus on genes that are under-studied in meiosis. To further prioritize the candidate genes, we evaluated the top VAAST genes with an assigned p-value < 0.05 (n = 404 genes, Table S2) for enriched gene ontology (GO) terms using the online tool ConsensusPathDB (CPDB). The GO cellular compartment (CC) terms such as “microtubule” (19 genes) and “kinesin complex” (5 genes) were the two most statistically significantly enriched GO CC terms (q < 0.05). All 5 genes from ‘kinesin complex’ were amongst the 19 ‘microtubule’ genes (*KIF18A, KIF5C, KIF16B, KIF20A, KLC1*) (Fig. 1B). See Table S3 for a complete list of gene lists and associated terms.

To further explore the interaction among the candidate genes, we constructed a protein-protein interaction (PPI) network using information from three common databases (CPDB, STRING, and GIANT). Among the 404 genes, 107 formed a connected network (Fig. S1, Table S4), including several genes that have an association with female aneuploidy (Known_genes, blue, Fig. S1), or predicted to be associated with meiosis in human by Meiosis Online (MO_human, orange, Fig. S1), including *KIF18A, FANCA, and KAT6A*. To further identify biologically relevant functional units, we applied the PAPER algorithm (*28*), which identifies root nodes and functional modules within a network. With this method, we identified seven discrete modules within the PPI network (Fig. 1C) and we identified GO terms enriched in these modules (Table S5). Within module 2, five genes (*KIF20A, CIT, ANLN, NCAPG2, KIF18A*) belong to the mitotic cell cycle process group (GO:1903047). Two of the five genes, *KIF18A* and *KIF20A*, are members of the kinesin superfamily of proteins (KIFs) identified in the VAAST-GO pipeline. We prioritized these two genes for validation studies because they overlapped between the VAAST-GO and the PPI approaches described here (also illustrated in Fig. 1B). Prior to conducting validation studies, we confirmed that these kinesins are expressed in mouse oocytes. Previous studies indicated KIF20A spindle localization in mouse and porcine oocytes (*29, 30*) which we verified here (Fig. S2A). We found that KIF18A also localized to the Metaphase I spindle, consistent with previous reports in mitotic and meiotic cells (*23, 31–33*) (Fig. S2B).

### Motor domain inhibition in candidate kinesins increases egg aneuploidy

After identifying four variants in *KIF20A* and two in *KIF18A* that are enriched in EEA patients compared to low-aneuploidy controls, we sought to prioritize these variants for experimental validation based on their potential functional significance (Fig. 1D). Both KIF genes have motor domains, the enzymatic regions with dynamic microtubule-binding affinity and ATP-hydrolysis activity that confer motility to the protein. Each gene contained EEA patient variants within this domain (Fig. 1D). To begin to test the hypothesis that the kinesin motor domain mutations could cause EEA, we first determined whether kinesin motor function is important for euploidy. Therefore, we assessed whether biochemical inhibition of each candidate kinesin’s motor domain causes egg aneuploidy (Fig. 2A). We treated Prophase I oocytes with either Sovilnesib to inhibit KIF18A motor function (*34*) or Paprotrain to inhibit KIF20A motor function (*35*). We then cultured the oocytes to metaphase II and performed an *in-situ* chromosome counting assay to evaluate whether each egg was aneuploid or euploid. We found that the frequency of aneuploidy increased significantly in oocytes treated with either Paprotrain or Sovilnesib. Specifically, 52.3% of oocytes were aneuploid in the Paprotrain-treated group, and 40.0% of oocytes were aneuploid in the Sovilnesib-treated group, compared with 15.9% and 20.8% aneuploidy in oocytes treated with vehicle controls, respectively (Fig. 2B,C). These results indicate that kinesin motor activity is required for egg euploidy and that impairment of this function can lead to egg aneuploidy.

**Figure 2.**
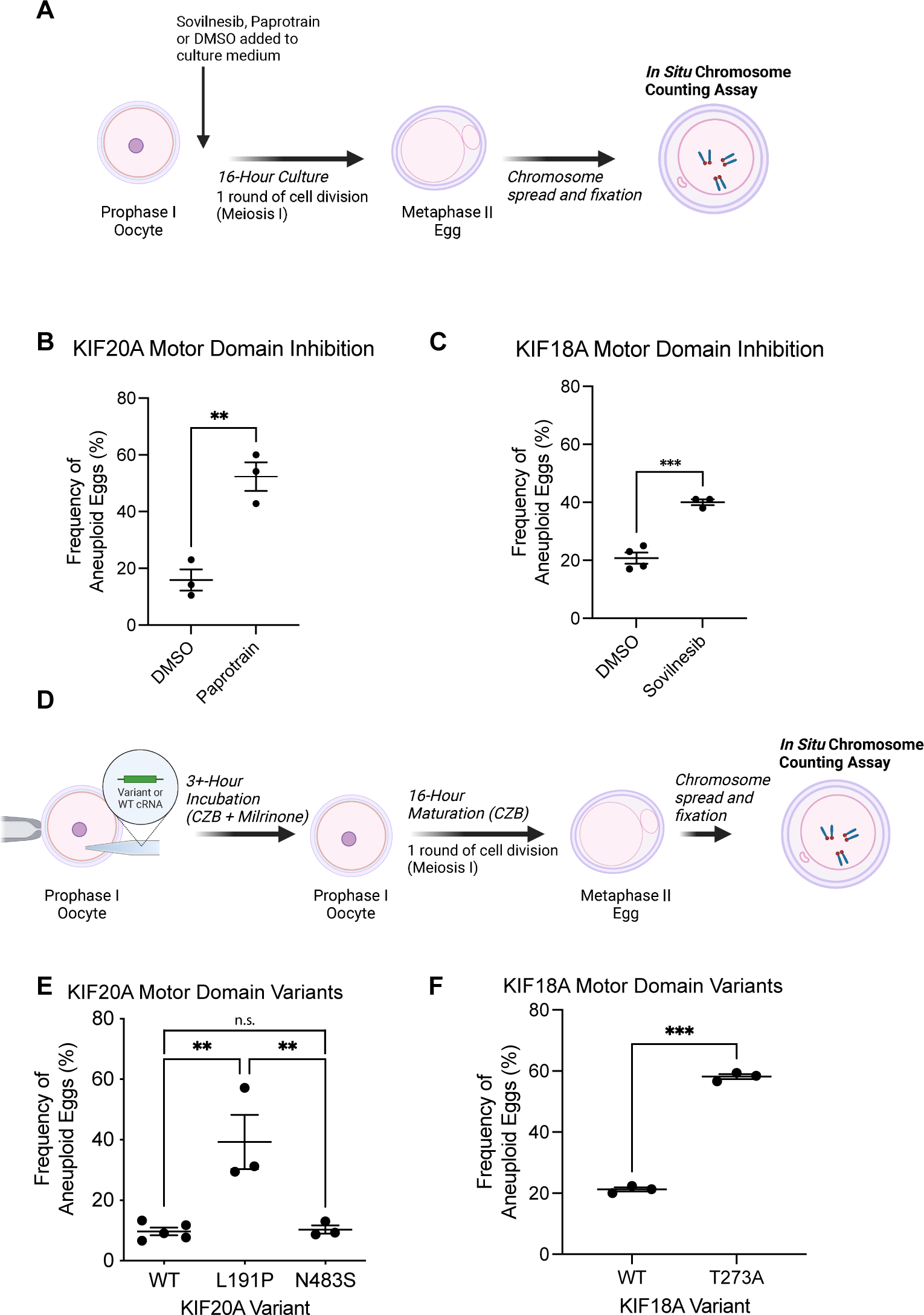
Evaluation of kinesin motor domains in mouse oocytes. (A) Diagram of strategy to chemically inhibit KIF18A (Sovilnesib) or KIF20A (Paprotrain) and assess frequency of aneuploidy. (B) Frequency of aneuploidy in oocytes treated to inhibit KIF20A. DMSO, n = 94 oocytes from 3 mice; 15 μM, n = 120 oocytes from 3 mice. Two-tailed t-test. p = 0.0043. (C) Frequency of aneuploidy in oocytes treated to inhibit KIF18A. DMSO, n = 58 from 5 mice; 500 nM, n = 67 from 3 mice. Two-tailed t-test. p = 0.0005. (D) Diagram of strategy to express motor domain variants and assess frequency of aneuploidy. (E) Frequency of aneuploidy in oocytes overexpressing *KIF20A* patient variants. One-way ANOVA. WT, n = 39 oocytes from 3 mice; L191P, n = 41 oocytes from 3 mice; N483S, n = 38 oocytes from 3 mice. WT v L191P, p = 0.0026; WT v N483S, p = 0.0058. (F) Frequency of aneuploidy in oocytes overexpressing *KIF18A* variants. Two-tailed t-test. p < 0.001. WT, n = 37 from 3 mice; T273A, n = 35 from 3 mice. Data are represented as mean ± SEM. *p < 0.05, ** p < 0.01, *** p < 0.001, **** p < 0.0001.

### Kinesin variants enriched in EEA patients increase egg aneuploidy

Having determined that the kinesin motor domain is required for egg euploidy, we turned our attention to the three kinesin motor domain variants enriched in our EEA patient cohort (Fig. 1D). To test the hypothesis that these variants cause EEA by increasing egg aneuploidy, we evaluated each variant for its ability to cause egg aneuploidy when overexpressed in mouse oocytes. We microinjected *Gfp*-tagged cRNA encoding each variant into age-matched, wild-type mouse oocytes arrested in Prophase I, cultured the oocytes *in vitro* to Metaphase of meiosis II (Met II), and performed an *in situ* chromosome counting assay to determine whether each cell was aneuploid (Fig. 2D).

Exogenously expressed KIF18A and KIF20A variants localized in the same fashion as the reference alleles (compare Fig. 3B,E to S2A,B). Exogenous KIF20A-GFP localized to Telophase I midbodies (Fig. 3B), as reported of endogenous KIF20A in mouse oocytes (*36*). KIF18A-GFP localized similarly to endogenous KIF18A at the Metaphase I spindle (compare Fig. 3E to Fig. S2B). Of note, KIF18A-GFP demonstrated pronounced accumulation at the kinetochore-fiber tips, similar to a recent report using a transgenic KIF18A-GFP mouse strain (*37*) (Fig. 3E) and consistent with somatic cells’ localization of KIF18A (*31*). Therefore, the GFP tag does not alter subcellular localization. Furthermore, these data suggest that exogenous hKIF20A-GFP, hKIF18A-GFP, and the patient variants therein are similarly competent as the endogenous protein in their ability to bind to microtubules. Further, the localization at plus end tips suggests that hKIF18A-GFP and the tested motor domain variants (Fig. 1D) are competent to conduct plus-end-directed motor activity and localization of hKIF20A-Gfp at midbodies are competent to move CPC cargo during Telophase I.

**Figure 3.**
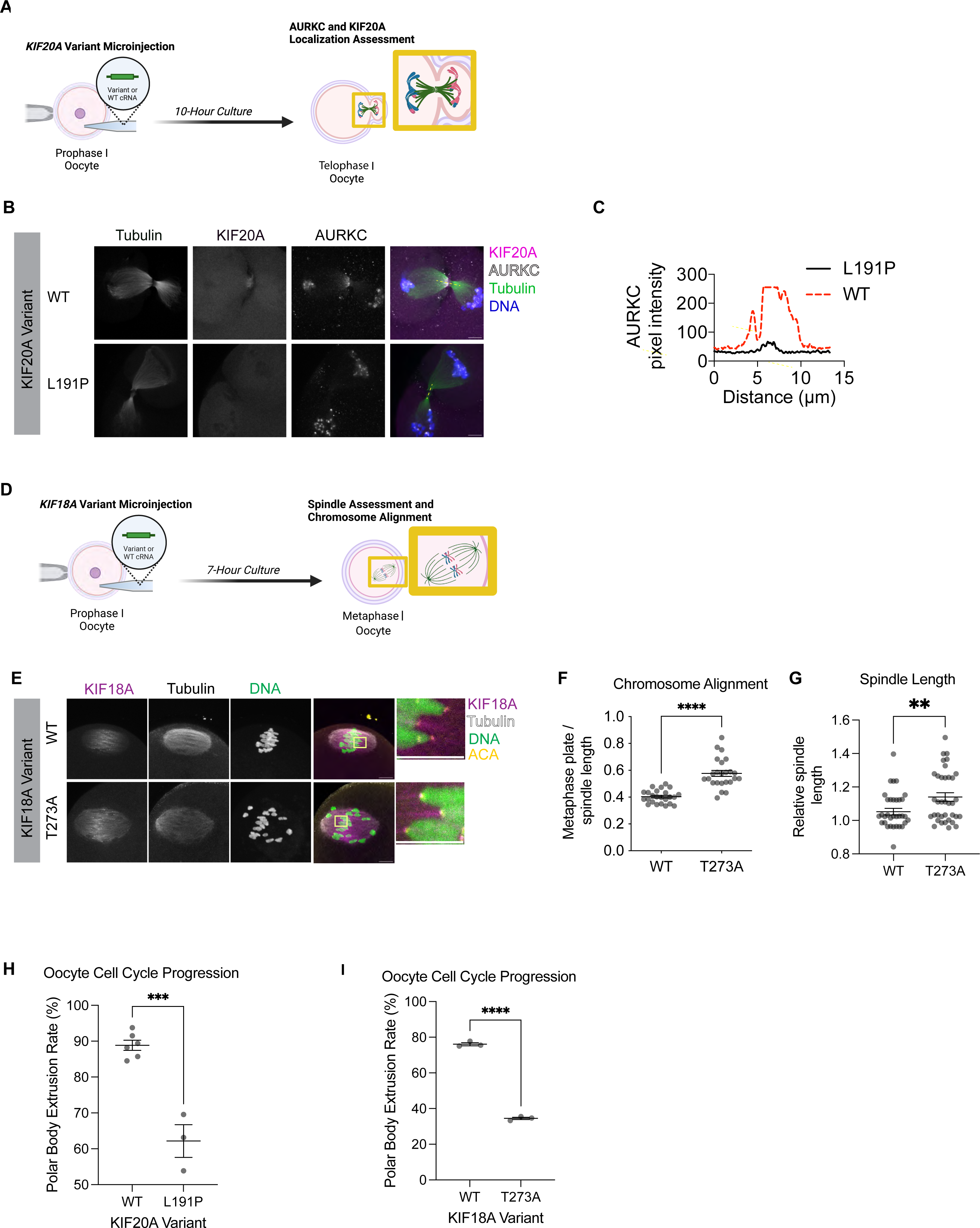
Functional KIF20A and KIF18A assays in oocytes expressing patient variants. (A) Diagram of strategy to assess Telophase I AURKC localization. (B-C) AURKC signal at the midzone in oocytes expressing variant KIF20A. Oocytes were fixed and immunolabeled for AURKC (gray), α-tubulin (green) and DNA (blue). GFP-hKIF20A, magenta. n > 15 oocytes/group. (C) Intensity of AURKC along yellow line in (B). (D) Diagram of strategy to assess Metaphase I spindles in oocytes expressing variant KIF18A. (E) Metaphase I chromosome alignment images. α-tubulin (gray), ACA (yellow), and DNA (green), hKIF18A (magenta). Zoom-insets of k-fiber tips on right. (F-G) Quantifications from E. WT, n = 34 oocytes; T273A, n = 36 oocytes. Each dot represents one oocyte. Student’s t-test. F: p *=* 0.0003. G: p = 0.009. (H-I) Frequency of polar body extrusion in oocytes overexpressing *KIF20A* and *KIF18A* variants. Dots represent one replicate. H: WT, n = 94 oocytes; L191 *n* = 92 oocytes. Student’s t-test. *P =* 0.002. I: WT, n = 81 oocytes; T273A, n = 89 oocytes. Student’s t-test. p < 0.0001. Data are represented as mean ± SEM. ns, not significant; * p < 0.05, ** p < 0.01, *** p < 0.001, **** p < 0.0001. Scale bars 5μm.

Upon performing the chromosome-counting assay, we observed that oocytes expressing either KIF20A^L191P^ or KIF18A^T273A^ had a significantly higher frequency of egg aneuploidy compared to controls (Fig. 2E,F). However, expression of the KIF20A^N483S^ motor domain variant did not significantly change aneuploidy frequency relative to controls (Fig. 2E). As a control, we confirmed that the exogenous WT and variant proteins were expressed at a similar level by assessing GFP levels by Western blotting (Fig. S3). Having determined that KIF20A^L191P^ and KIF18A^T273A^ increase egg aneuploidy, we next sought to understand the functional impact of these patient kinesin motor domain variants on eggs.

### KIF20A^L191P^ and KIF18A^T273A^ disturb critical processes in oocyte meiosis I

Meiosis I is the process by which oocytes prepare to segregate their chromosomes; most egg and embryo aneuploidies arise from errors in meiosis I (*1–3*). Therefore, to evaluate the functional consequence of each EEA-patient-enriched kinesin motor domain variant on egg quality, we assessed the effect of exogenous expression of the EEA-patient-enriched motor domain variants on meiosis I in detail. We used gene-specific assays to assess the impact of the aneuploidy-causing variants on KIF20A and KIF18A function.

In mitotic cell division, KIF20A binds to and transports the CPC from chromosomes to the spindle midzone during anaphase where the complex then promotes formation of the cleavage furrow and regulates cytokinesis (*17–20*). This localization is critical for the recognition of lagging chromosomes in anaphase and telophase to delay cell division (*21*). In these cells, depletion of KIF20A prevents CPC localization to the spindle midzone and midbody (*38*). In meiotic cells, like mouse oocytes, Aurora kinase C (AURKC) is the catalytic subunit of the CPC (*39, 40*). To assess whether this function is maintained in oocytes expressing KIF20A^L191P^, we next evaluated the effect that KIF20A^L191P^ had on AURKC/CPC localization at Telophase I. We microinjected oocytes with either *Gfp*-tagged WT*-hKIF20A* or h*KIF20A^L191P^*, cultured them for 10 hours, fixed them, and labeled for tubulin, to reveal the midbody, and AURKC (Fig. 3A).

Oocytes expressing the reference allele (WT) had enriched AURKC in the meiotic midbody at Telophase I (Fig. 3B, top). The exogenous hKIF20A co-localized with AURKC in this region (Fig. 3B, merge). In contrast, AURKC did not localize to meiotic midbodies and instead remained on chromosomes in oocytes expressing hKIF20A^L191P^ (Fig. 3B, bottom, 3C). The failure of the CPC to re-localize was accompanied by loss of hKIF20A^L191P^ localization in this region. We confirmed this phenotype by assessing AURKC meiotic midbody localization in Paprotrain-treated (KIF20A-inhibited) oocytes. AURKC midbody signal decreased in oocytes cultured in 15 μM Paprotrain as compared to vehicle control (Fig. S4). Paprotrain inhibits KIF20A’s ATPase function (*35*). These data indicate that hKIF20A^L191P^ fails to migrate to the midbody along with its cargo, the CPC, and suggest that hKIF20A^L191P^ has altered motor function.

We next assessed KIF18A function in oocytes expressing hKIF18A^T273A^. KIF18A is a motor protein that promotes chromosome alignment by dampening chromosome oscillation (*23, 41*). Loss of KIF18A causes chromosome misalignment in mitotic cells, so we assessed this function in oocytes expressing hKIF18A^T273A^. We microinjected mouse oocytes with cRNA encoding hKIF18A^T273A^, matured the cells to Metaphase I, and labeled spindles and chromosomes (Fig. 3D). Then, we calculated the ratio between the length of the metaphase plate (the distance between the two farthest chromosomes) and spindle length, as a metric of chromosome alignment. Compared to oocytes expressing the reference allele (WT), we observed an increase in the frequency of chromosome misalignment when oocytes expressed KIF18A^T273A^ (Fig. 3E,F).

Notably, we also found an increase in spindle length (Fig. 3G), a phenotype that also occurs with KIF18A-knockdown (*23, 31*). KIF18A is a microtubule-stabilizing enzyme and its inactivity in meiotic and mitotic cells is frequently associated with abnormally long spindles (*23, 31, 33, 42*). Together, these results suggest that KIF18A function is altered in oocytes expressing hKIF18A^T273A^.

Errors in meiosis I can lead to meiotic delay or arrest through activation of cell cycle checkpoints (*43–45*). Therefore, we assessed meiotic progression, as measured by the frequency of polar body extrusion (PBE) after 16 hours of culture, in oocytes expressing hKIF20A^L191P^ or hKIF18A^T273A^. We found a significant reduction in meiotic progression in oocytes expressing hKIF20A^L191P^ (62% PBE vs 89% in WT control; Fig. 3H) and hKIF18A^T273A^ (35% PBE vs 76% in WT control; Fig. 3I). These results suggest that although some proportion of oocytes expressing kinesin motor domain variants enriched in EEA patients progress through meiosis I to yield aneuploid eggs, another population of these oocytes fails to proceed through cytokinesis in a timely fashion, likely due to errors in meiosis I.

### mKIF18A^S273A^ accelerates female reproductive aging by prematurely increasing egg aneuploidy

Having validated the causality of hKIF20A^L191P^ and hKIF18A^T273A^ in egg aneuploidy *in vitro*, we considered the utility of generating pre-clinical *in vivo* models of these variants. First, we assessed the allele frequency (AF) of these kinesin motor domain variants in EEA patients as compared with patients with low rates of embryonic aneuploidy.

KIF20A^L191P^ allele was present in 1 of 178 patients with an AF of ∼0.0028. In gnomAD, this allele is rare (AF 0.00001533 in the European non-Finnish population). Because this allele is very rare in the general population and therefore has restricted impact potential, we elected not to generate a mouse model of this variant. In contrast, we found that hKIF18A^T273A^ was present in 17 out of 178 patients with an AF of ∼0.05 (Fig. 1A, blue dots). The same allele was found at a similarly high AF in the gnomAD database for the European non-Finnish population (0.03881), indicating this is a relatively common variant. In our cohort, hKIF18A^T273A^ heterozygosity is strongly associated with the high-rate group (13 HRG patients, and only 3 LRG patients), and one patient, a 25-year-old with 45% aneuploidy, was homozygous (Fig. 1A, black dot).

Having established that h*KIF18A^T273A^* is a common allele that is both enriched in EEA patients and functionally pathologic to mouse eggs *in vitro*, we sought to establish a pre-clinical *in vivo* model of h*KIF18A^T273A^*. We generated a knock-in mouse model by using CRISPR/Cas9 genome editing to replace the mouse codon TCA that encodes Serine 273 in *Kif18a* with a CGC encoding Alanine (Fig. S5). This mouse model provided two opportunities: (1) validate the variant as a genetic cause of EEA in an *in vivo*, mammalian context; (2) assess the functional impact of the variant on whole-organism fertility and endogenously expressing mammalian eggs.

To evaluate the consequence of harboring the KIF18A^S273A^-encoding allele on female fecundity, we performed fertility trials in which female mice of each *Kif18a* genotype were paired with highly fertile B6D2/J males for six months and assessed for litter size over time. At reproductively young ages (∼2 months of age), WT, heterozygous, and homozygous mutant dams were similarly fertile (p > 0.9999; Fig. 4A-B). However, after five months of age, the litter sizes of homozygous mutant dams were significantly smaller, compared with WT dams (p = 0.0425). As a result, over the whole trial, homozygous mutant mice had significantly different fertility from WT mice (p = 0.0009; Fig. 4A). We concluded that the female subfertility phenotype of KIF18A^S273A^- encoding mice arises only in the context of moderately increased maternal age, suggesting an acceleration of the typical murine age-dependent decline in fertility (*46*).

**Figure 4.**
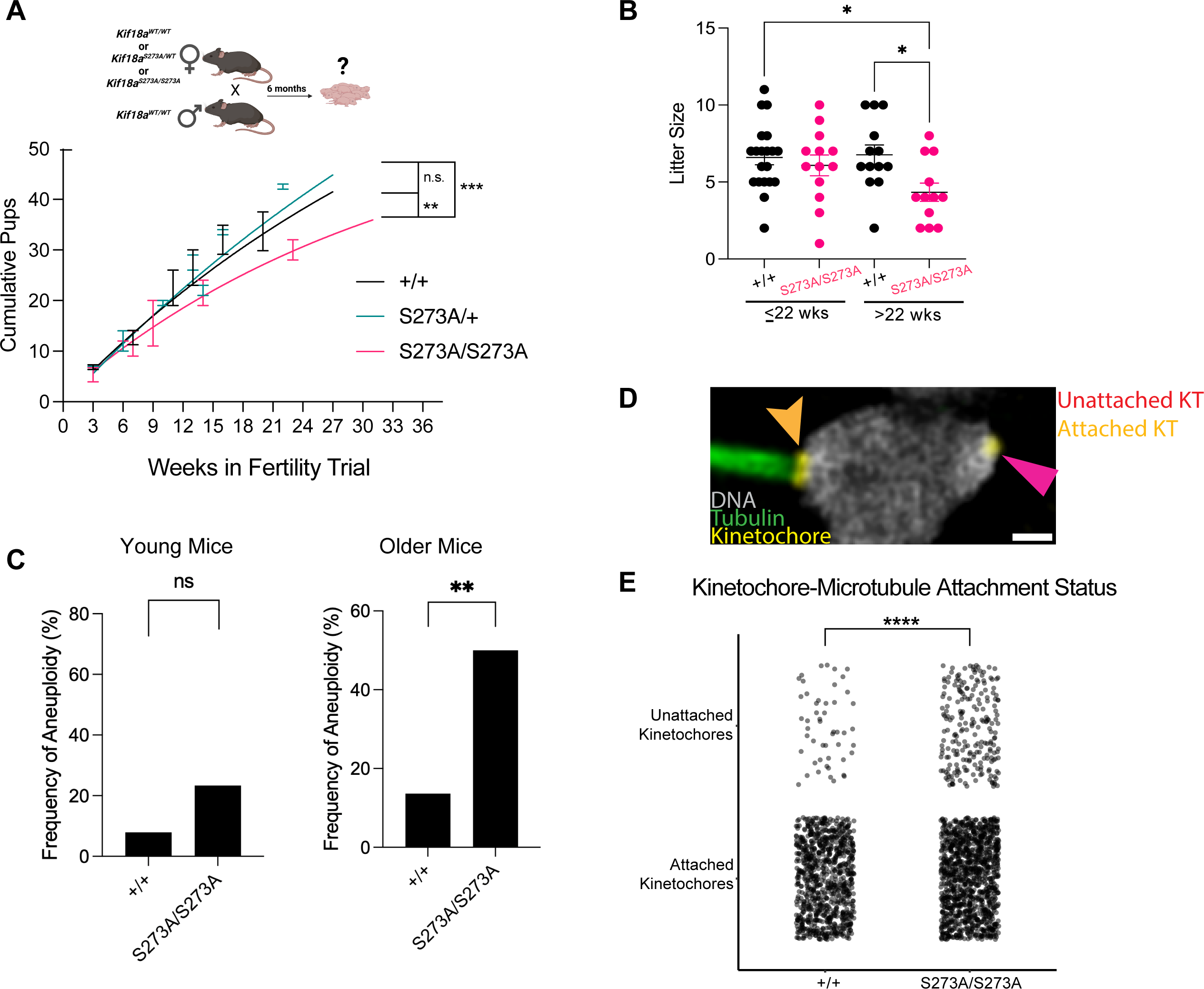
Female *Kif18a*^S273A^ mice experience accelerated reproductive aging. (A) Cumulative pups from fertility trial. +/+ vs S273A/S273A (p = 0.0009). +/+, n = 5; S273A/+, n = 4; S273A/S273A, n = 4. Comparison of fits test p = 0.0001. Multiple comparisons: +/+ vs. S273A/+, p = 0.706; +/+ vs. S273A/S273A, p = 0.0009; S273A/+ vs. S273A/S273A, p < 0.0001. (B) Litter size analysis of young (≤ 22 weeks) and older mice (> 22 weeks). ANOVA, Tukey’s multiple comparisons test. p = 0.030; +/+ young vs. S273A/S273A older p = 0.0371; +/+ older vs. S273A/S273A older p = 0.0425. (C) Frequency of aneuploid eggs in young (8-10 weeks) and older (> 20 weeks). 5 replicates. Younger: +/+, n = 38; S273A/S273A, n = 60; p = 0.059. Older: +/+, n = 22 oocytes; S273A/S273A, n = 30; p = 0.008. Data from each genotype pooled; Fisher’s exact test. (D) Bivalent with microtubule-attached (pink arrowhead) and -unattached kinetochores (orange arrowhead). DNA (gray), alpha-tubulin (green), anti-centromere antibody (yellow). (E) Quantification of unattached and attached kinetochores. Dots represent single kinetochores. All kinetochores are pooled. +/+, n = 916; S273A/S273A, n = 1,370. Chi-square analysis. p < 0.0001. Data are mean ± SEM. ns, not significantly different; * p < 0.05; ** p < 0.01, *** p < 0.001.

It is likely that the effect of the variant is sex- and meiosis-specific. Litter sizes were not significantly different between *Kif18a* variant males (p = 0.065; Fig. S6), indicating that the effect of *Kif18a^S273A^* on fertility is specific to females. In addition, mitotic chromosome segregation was unaffected by *KIF18A^T273A^*when expressed in somatic U2OS cells, both in terms of chromosome alignment (p = 0.559; Fig. S7A,B) and chromosome segregation errors (p = 0.465; Fig. S7C). hKIF18A^T273A^ accumulated at the plus-end tips of microtubules throughout mitosis in U2OS cells, with a statistically significant but modest 6% decrease in accumulation compared to hKIF18A^WT^ (*p* = 0.0193, T273A 0.840 v. WT 0.893; Fig. S7D-E). Further studies examining the relationship between *KIF18A^T273A^*, sex, aging, and other body systems are required to fully elucidate the consequences of the variant on both male mammals and somatic tissues.

Having established that *Kif18a^S273A^* exacerbates the age-dependent decline in female fertility, we next asked whether egg aneuploidy caused this decline. We assessed egg aneuploidy at the age at which fertility was similar (∼9 weeks of age, young) and at the age at which inter-genotype fertility diverged (∼20 weeks of age, older). In young mice, the proportion of aneuploid eggs trended upward in homozygous mutant mice but was not significantly different from WT mice (Fig. 4C; 23% vs 8%; p = 0.059). In older mice, aneuploidy was significantly higher in homozygotes, compared with controls (Fig. 4C; 50% vs 13%; p = 0.008). To identify the mechanism by which *Kif18a^S273A^* causes egg aneuploidy, we examined kinetochore-microtubule attachments, a key cause of aneuploidy, in oocytes collected from older (20-week-old) *Kif18a^S273A/S273A^* mice. We treated these oocytes with a brief incubation in ice-cold media to depolymerize microtubules not stably attached to kinetochores. We analyzed ∼1,000 kinetochores per genotype and found a significant increase in unattached kinetochores in oocytes from aged *Kif18a^S273A^* mice compared to age-matched WT controls (Fig. 4D-E; 16.1% vs 5.5% p < 0.0001). This defect may result from altered KIF18A-mediated microtubule dynamics at the microtubule K-fiber tip.

Collectively, these data demonstrate that *KIF18A^S273A^* specifically increases egg aneuploidy and that the effect of KIF18A^S273A^ on egg aneuploidy is compounded by maternal age-dependent changes in the egg. Altogether, the data support a model in which KIF18A^S273A^ migrates to the k-fiber tip, but fails to regulate microtubule dynamics, resulting in elongated spindles, misaligned chromosomes, and unattached kinetochores. Furthermore, these data indicate that these changes lead to prematurely increased egg aneuploidy, translating finally to an accelerated decline in female fertility, suggesting a new inherited genetic mechanism of female infertility via the kinesin motor domain.

## Discussion

In this study, we employed a bioinformatic strategy to identify plausible common pathological variants of accelerated loss of egg quality. To our knowledge, this is the first report implicating kinesin motor domain variation, and kinesin variants in general, as a causal genetic mechanism in female infertility patients. Notably, we provide the first *in vivo*, preclinical validation of a human genetic variant for the trait of EEA in female IVF patients. These findings regarding a key mechanism of female fertility and pregnancy loss are particularly timely, given the changing regulatory context of female fertility, the increase in maternal age at first pregnancy (from 21.4y in 1970 to 27.3y in 2021) (*47, 48*), and the rapidly expanding use of assisted reproductive technologies, resulting in 3% of all US infants (*49*).

Genetic variation may explain some of the heterogeneity in the relationship between egg aneuploidy and maternal age: genetic variants have been identified in multiple genes that may influence egg and embryonic aneuploidy at younger-than-average ages (*50, 51*). For example, one genetic variant in *CEP120* was identified in individuals with higher-than-average aneuploidy and its function was demonstrated in an *in vitro* mouse oocyte system (*52*). However, to date, no aneuploidy-causing variant has been biologically validated in a whole-organism model. The organism-level validation is important because another study of oocyte meiosis genes demonstrates that *in vitro* perturbations may not translate to *in vivo* pathogenicity (*53*). Furthermore, no human genetic variants have yet been identified to cause EEA specifically via accelerated maternal aging, as we demonstrate here. As a result, no predictive genetic biomarkers of EEA exist. One major obstacle to the identification of EEA variants is the paucity of genotype-phenotype data available, resulting in relatively small patient cohorts. Functional biological validation of candidates thus is a “gold standard” to identify true genetic causality in EEA.

We found that kinesin motor domain variants in *KIF20A* and *KIF18A* identified in EEA patients had a deleterious effect on mouse oocytes via our overexpression bioassay, causing aneuploidy and abrogating gene-specific functions required for correct chromosome segregation (Fig. 5). When we knocked the homologous variant of KIF18A^T273A^ into the endogenous *Kif18a* locus in mouse, we found that homozygous mutants had increased egg aneuploidy and decreased fertility at moderately advanced maternal age. T273 resides in the KIF18A motor domain, specifically in the Loop 11/Switch II region of the kinesin and resides at the boundary of the α4 helix (*54–56*).

**Figure 5.**
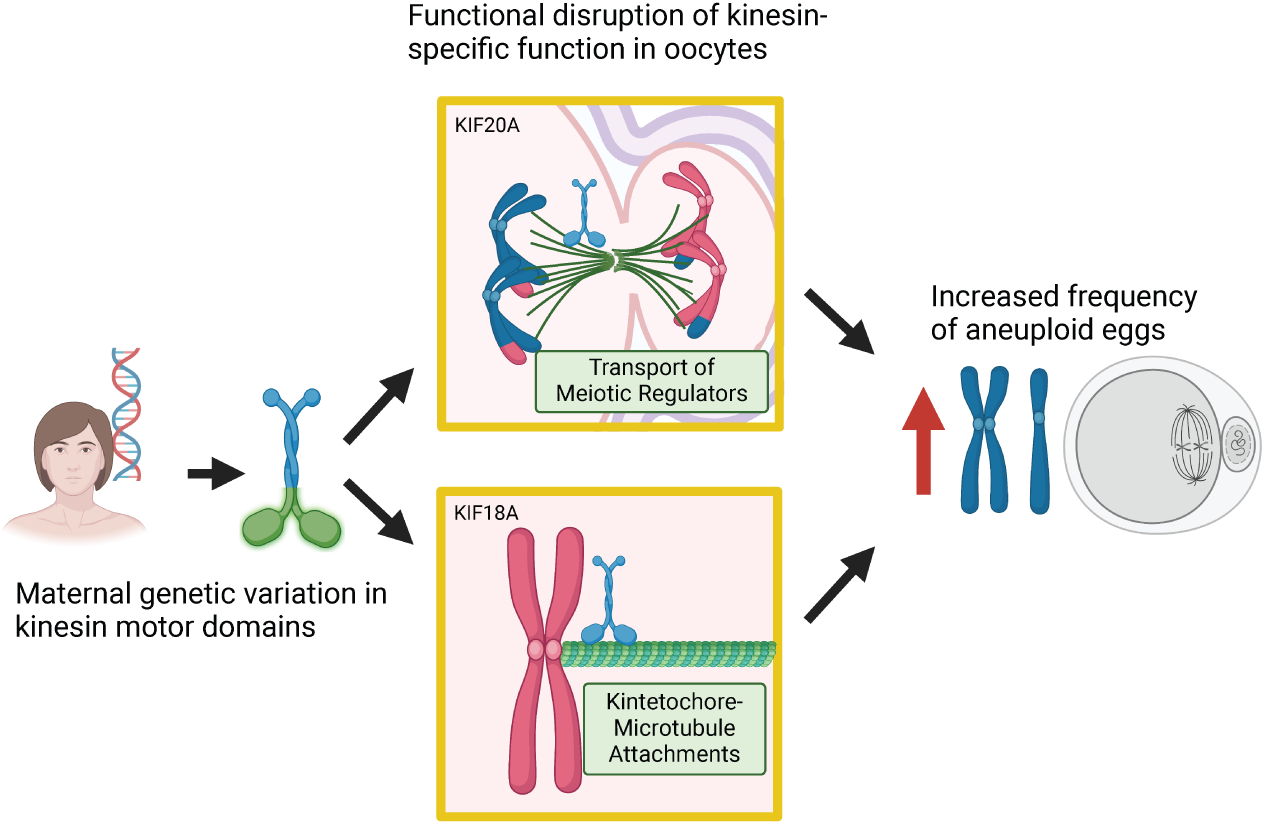
Summary schematic of overall findings.

The α4 helix region is implicated in tubulin binding. Phosphorylation of nearby S284 alters motor directionality causing chromosome misalignment and changes in spindle length (*55*). Although we do not know if T273 is a phospho-acceptor site, our data are consistent with altered function when a residue in this critical region is altered.

A recent report asserted that KIF18A is nonessential in meiosis I based on a ZP3-Cre *Kif18a* knockout (*37*), a statement seemingly in conflict with the results shown here. However, the experiments shown here found an effect with age. In contrast, the ZP3-Cre *Kif18a* knockout was only evaluated in young mice. When considered in this light, our work aligns with and stretches beyond the existing literature. Homozygous mutant mice did not show subfertility until they approached ∼20 weeks of age, which is biologically akin to a human’s late twenties and early thirties (*57*) and parallels the human patient data (Fig. 1A). Our data suggest that this variant is pathogenic for EEA only when coupled with a “second hit” of normal mechanisms of aging, revealing a complex genotype-phenotype relationship. It is well established that cohesin, the molecular “glue”, is lost progressively from chromosomes in aging human and mouse oocytes, resulting in age-dependent increases in aneuploidy (*4–7*). Further experimental analysis would be required to elucidate the precise molecular interplay between maternal aging, KIF18A, and egg aneuploidy. Large-scale human trials are required to ultimately determine the diagnostic utility of *KIF18A^T273A^* and other kinesin motor domain variants.

Our study has several limitations. The sample size of our patient cohort is relatively small, limiting our power to identify variants and genes contributing to the aneuploidy risk. To address this limitation, we developed an enhanced variant prioritization pipeline and performed deep biological validation, both *in vitro* and *in vivo*. Another limitation is that our analysis focuses on women with primarily European ancestry, which is due to the overrepresentation of this group in the sampled fertility clinic patient populations. Therefore, our results may not generalize to other ethnic groups. New genomic biobanks of egg aneuploidy phenotypes and future analyses of ultra-low-coverage genetic data from PGT-A should aim to include other ethnic groups (*58*). A third limitation is the use of mouse oocytes, rather than human oocytes, to biologically validate our results. Currently, the only human oocytes available for use in laboratories are those that IVF clinics have discarded due to slow or arrested cell cycle progression; these oocytes would be a highly genetically heterogenous and functionally nebulous system in which to evaluate the consequence of specific genetic variants on oocyte meiosis. Mouse oocytes undergo similar meiotic maturation steps and can be genetically and environmentally controlled. Therefore, the mouse oocyte system represents the best alternative approach.

This study lays the groundwork for future identification of EEA genetic variants by demonstrating an enhanced statistical and functional genetic pipeline (Fig. 1B) and identifies kinesin motor domains as a novel functional genetic hotspot for EEA (Fig. 5). Moreover, this study provides the deepest biological validation of a candidate clinical variant for causation of EEA to date and the first *in vivo* mouse model of an EEA patient variant, paving the way for biomarker development. Identification of precision genetic biomarkers for EEA, could provide a personalized approach to fertility care, bringing a genomic medicine lens to fertility treatment.

## Material and Methods

### Ethical approval, sample selection, and whole-exome sequencing dataset

Patient DNA samples were obtained from Reproductive Medicine Associates of New Jersey (RMANJ) DNA Bank, and the project was approved by IRB #RMA1-09-165 at Copernicus Group IRB and IRB #Pro2018000106 at Rutgers University. Preimplantation genetic testing for aneuploidy (PGT-A) was performed on biopsies of trophectoderm cells from day 5 human blastocysts. Aneuploidy rate was calculated for “White, non-Hispanic” patients with a minimum of four embryos tested. Samples with extreme aneuploidy rates were selected using a robust nonlinear regression analysis as previously described (*25*).

Whole-exome sequencing libraries were constructed by Novogene (Sacramento, CA, USA) from maternal peripheral blood DNA samples using the Agilent SureSelect Human All Exon V6 kit by Agilent Technologies (Santa Clara, CA, USA). The libraries were sequenced on the Illumina sequencing platform using the 2×150 base-pairs paired-end format (Illumina, San Diego, CA, USA). Data was aligned to the human reference genome (hg19) using BWA (*59*) and the joint genotyping was performed using the GATK v3.8 pipeline following the GATK best practices (*60*).

### Gene prioritization with VAAST

Variant Annotation, Analysis and Search Tool (VAAST) (*26*) was applied to identify candidate genes associated with high aneuploidy risk, as previously described (*52*). VAAST was ran with the following parameters:

$ VAAST --lh y -r 0.05 -d 1e5 -o outputfile -m lrt -k ref_gff controls.cdr cases.cdr

### GO and protein-protein interaction network analyses

GO enrichment analysis was performed using the over-representation analysis tool on CPDB (http://cpdb.molgen.mpg.de/) (*61*) on candidate genes with p-values smaller than 0.05 in the VAAST analysis. To investigate protein–protein interaction (PPI) networks among VAAST genes, three databases were used: CPDB (*61*), STRING (*62*), and GIANT_v2 (*63, 64*). All types of interactions from the three databases were combined and filtered for the network construction, as previously described (*51, 65*). The network was constructed using a customized Python code (https://github.com/JXing-Lab/network-ppi).

Modules within the PPI network were identified using PAPER (*28*), a program which employs the *Preferential Attachment Plus Erdős–Rényi* model for community detection within a network. PAPER was run on the original PPI network using the following parameters: method=collapsed; Burn=1000; M=5000; size_thresh=0.02; birth_thresh=0.6. EMP (https://github.com/Shamir-Lab/EMP) was used to conduct the GO enrichment analysis on each discovered module and to calculate its empirical p-value (*66*).

### Validation of the variants

Seven candidate variants in *KIF18A and KIF20A* were PCR amplified and the amplicons were sequenced using Illumina Nextseq (San Diego, CA, USA) at the Foundation for Embryonic Competence (Table S6). All variants except one (*KIF20A:* chr5:137522092_C_T in hg19 coordinates) were validated. Therefore, *KIF20A:* chr5:137522092_C_T was excluded from the analyses.

### Mice

Sexually mature CF-1 female mice (6–10 weeks of age, Envigo (Indianapolis, IN, USA)) were used in this study, with the exception of the *Kif18a* knock-in mice. For *Kif18a* knock-in mice, *Kif18a* variant alleles were generated using CRISPR-Cas9 genome editing through Rutgers Genome Editing Shared Resource (New Brunswick, NJ, USA). C57BL/6J zygotes were microinjected with a mixture containing Cas9 protein (IDT, San Diego, CA, USA), an sgRNA (MilliporeSigma, Burlington, MA, USA) and an ssODN (IDT, San Diego, CA, USA), which contained homology arms and the S273A mutation (Table S7). Ear biopsies from founders were screened by PCR at the mutation site and the PCR products were digested with restriction enzyme *Hha I* (introduced by S273A and PAM change) to select founders with the homologous S273A change. PCR products from the selected founders were then subjected to next-generation-sequencing using Amplicon EZ kit by Azenta Life Sciences (Burlington, MA, USA) to confirm the S273A change. Homozygous founder mice (F0) were confirmed using Sanger sequencing. Founders were backcrossed to C57B6/J mice from Jackson Laboratory (Bar Harbor, ME, USA) for two generations and progeny were screened using PCR primers *KIF18AA* and *KIF18AB* (Table S7) by the gain of *Hha* I restriction site or by Sanger sequencing of PCR products. Then, animals were maintained in a C57B6/J background.

Housing and breeding use a 12 h light/12 h dark cycle and constant temperature was performed in the animal facility at Rutgers, the State University of New Jersey.

Food and water were provided *ad libitum*. All animal experiments performed in this study were approved by the Rutgers IACUC (protocol #201702497) and followed the guidelines set by the National Institutes of Health.

### Oocyte collection and *in vitro* maturation

Prophase I oocytes were collected as described (*67*) in minimum essential medium (MEM; #M0268, MilliporeSigma, Burlington, MA, USA) containing 2.5 μM milrinone (#M4659, MilliporeSigma, Burlington, MA, USA) to prevent the spontaneous meiotic resumption (*68*). Briefly, two days before oocyte collection, mice were primed with pregnant mare serum gonadotropin (PMSG, #493-10, Lee Biosolutions, Maryland Heights, MO, USA). After collection in MEM, oocytes were incubated in Chatot, Ziomek and Bavister (CZB) medium (*69*) without milrinone, in 5% CO2 and 37^°^C for the desired time of maturation depending on the meiotic stages to be evaluated (7 h for Metaphase I, 10 h for Telophase I and 16 h for Metaphase II).

### Drug treatments

Oocytes were cultured in CZB medium treated with either DMSO (control), 15 μM Paprotrain (#512533, Millipore-Sigma, Burlington, MA, USA) or 500 nM Sovilnesib (#TA9H98DB1F0F, Millipore-Sigma, Burlington, MA, USA). To assess maturation kinetics, oocytes were cultured in CZB after milrinone washout while in a EVOS M7000 Imaging System (Invitrogen, # AMF7000) at 37^°^C with 5% CO2. Oocytes were scored visually for nuclear envelope breakdown and polar body extrusion (PBE) as a proxy for metaphase II. To assess AURKC migration at Telophase I, oocytes were matured in CZB with the indicated dose of Paprotrain for 10 h (0 μM) or 14 h (15 μM) to adjust for the drug-dependent delay in maturation timing. To assess aneuploidy in oocytes treated with either Paprotrain or Sovilnesib, oocytes were matured in CZB with the indicated doses for 16 h. Subsequently, aneuploidy was assessed as described in the “*in situ* chromosome counting” section.

### Constructs and cRNA generation

To generate cRNA of *KIF20A* and *KIF18A* fused with enhanced green fluorescent protein (eGFP), the full-length coding sequences (CDS) of genes encoding human *KIF20A* (NM_005733.3) and *KIF18A* (NM_031217.4) were amplified from clones OHu24388 (Genscript, Piscataway, NJ, USA) and OHu28001D (Genscript, Piscatway, NJ, USA), respectively. The amplified CDSs were ligated into the *in vitro* transcription (pIVT) vector (*70*) containing eGFP fusion using the following primers:

*KIF20A*; For: 5′-ATGCGTCGACATGTCGCAAGGGATCCTTTCTC-3′, Rev: 5′-GGGCGTCTAGAGTACTTTTTGCCAAAAGGCC-3′

*KIF18A*; For: 5′-ATGCGTCGACATGTCTGTCACTGAGGAAGACCTG-3′, Rev: 5′-GCGGCGTCTAGATCTTAGATTTCCTTTTG-3′. All inserts were sequenced fully to ensure no PCR-induced mutations were introduced.

Mutagenesis was performed using the multi-site directed mutagenesis kit (#210515, Agilent Technologies, Santa Clara, CA, USA) according to manufacturer’s instructions. To mimic the human variants, pIVT-*KIF20A-GFP-WT* and *KIF18A-GFP-WT* plasmids were mutated using the following primers that contain the same variant nucleotide substitutions (in bold and underlined)

### Motor Domain Kinesin Variants

Kif20A p.Leu191Pro: 5′-GCTGATCTTCAATAGCC**C**CCAAGGCCAACTTCATC -3′

Kif20A p.Asn483Ser: 5′-AGGCCGTTCCTGCATGATTGTCA**G**TGTGAATCCCTG -3′

Kif18A p.Thr273Ala: 5′-TCCGGTGCTAAGGGG**G**CCCGATTTGTAGAAG -3′

### Non-Motor Domain Kinesin Variants (related to Figure S3)

Kif20A p.Ser44Phe: 5′-CCTGCTATCAGACTGCT**T**TGTCGTCTCTACCTC -3′

Kif20A p.Glu63Lys: 5′-TCCATCTGAGGACAGTATG**A**AGAAGGTGAAAGTATACTTG -3′

Kif18A p.Ser835Arg: 5′-AAAAGGAAACGGAAATTAACAAG**G**TCTACATCAAACAGTTCGTTAAC -3′

Mutations were confirmed by Sanger sequencing (Azenta, Piscataway, NJ, USA). *Kif20A* plasmids were linearized with *Ssp I* (#R0132S, New England Biolabs, Ipswich, MA, USA) and *Kif18A* plasmids with *Kas I* (#R0544S, New England Biolabs, Ipswich, MA, USA) restriction enzymes. pGEMHE-mEGFP-mTrim21 (#105519, Addgene, Watertown, MA, USA) plasmid was linearized with AscI (#R0558S, New England Biolabs, Ipswich, MA, USA). All linearized plasmids were then purified with QIAquick PCR Purification kit (#28104, Qiagen, Venlo, The Netherlands) and *in vitro* transcribed using a T7 mMessage mMachine Kit (#AM1345, Thermo Fisher Scientific, Waltham, MA, USA). The cRNAs were purified using an RNAeasy kit (#74104, Qiagen, Venlo, The Netherlands) and stored at −80°C.

### Microinjection

Prophase I-arrested oocytes were microinjected with the indicated cRNAs in MEM medium containing milrinone using a Xenoworks microinjector (Sutter Instruments, Novato, CA, USA) as described before (*71*). Then, oocytes were kept in milrinone-containing CZB medium for at least 2 h or overnight for protein expression before downstream assays.

### *In situ* chromosome counting

As described previously (*72*), after *in vitro* maturation of Prophase I oocytes for 16 h, Metaphase II eggs were incubated with 100 µM Monastrol (#M8515, MilliporeSigma, Burlington, MA, USA), dissolved in dimethyl sulfoxide (#472301, MilliporeSigma, Burlington, MA, USA) in CZB medium for 2 h before fixation in 2% paraformaldehyde (PFA, #P6148, MilliporeSigma, Burlington, MA, USA) in phosphate-buffered saline (PBS). Eggs were stained with Anti-Centromeric Antibody (IF: 1:30; #15-234, Antibodies Incorporated, Davis, CA, USA) to detect centromeres and mounted in VectaShield (#H- 1000, Vector Laboratories, Newark, CA, USA) with 4′, 6-Diamidino-2-Phenylindole, Dihydrochloride (DAPI; #D1306; 1:170, Life Technologies, Carlsbad, CA, USA). Eggs were imaged using a Leica SP8 confocal with 0.5-µm z-intervals. Chromosome counting was performed with NIH ImageJ software (*73*). Normal chromosome number for a mouse egg is 20 pairs of sister chromatids; an egg with any deviation from this number was considered aneuploid.

### Female Fertility Trials

Sexually-mature female mice (8 to 10 weeks old) bearing either *Kif18a^+/+^, Kif18a^S273A/+^,* or *Kif18a^S273A/S273A^*genotypes were housed in monogamous pairings with B6D2F1/J male mice (Jackson Laboratory, Bar Harbor, ME, USA). The animals remained paired for six months and were allowed to breed freely. Daily observations were made for pup counting starting at day 18 based on the mouse gestational period (18-21 days).

### Histology

Subsequent to each fertility trial, female mice harboring the *KIF18A* alleles were euthanized, and ovaries were collected for histology. Ovaries were fixed in Davidson’s modified fixative (overnight at 4^°^C), washed in 70% ethanol, embedded in paraffin, serially sectioned (5 μm), and stained with hematoxylin and eosin (H&E) (Rutgers Translational Sciences Research Pathology Services, Piscataway, NJ). Three ovarian sections per animal were evaluated for the presence of *corpora lutea* to confirm ovulation.

### Antibodies for Oocyte Immunofluorescence and Western Blotting

The following primary antibodies were used for immunofluorescence (IF) and Western blotting (WB): rabbit anti-KIF18A (IF:1:50, #19245-1-AP, ProteinTech, Rosemont, IL, USA), mouse anti-KIF20A (IF:1:100, #67190-1-Ig, ProteinTech, Rosemont, IL, USA), rabbit anti-GFP (WB:1:500, #G1544, MilliporeSigma, Burlington, MA, USA), rabbit anti-α-tubulin (WB: 1:1000; IF:1;50; #11H10, Cell Signaling Technology, Danvers, MA, USA), sheep anti-alpha/beta-tubulin (IF:1:100, #ATN02, Cytoskeleton, Denver, CO, USA), Rabbit anti-EEA1 (IF:1:100, #3288T, Cell Signaling Technology, Danvers, MA, USA), human ACA (IF: 1:30; #15-234, Antibodies Incorporated, Davis, CA, USA), rabbit anti-AURKC (IF: 1:200; #A400-023A-BL1217, Bethyl Laboratories, Montgomery, TX), mouse anti-acetylated α-tubulin (IF: 1:100, #T7451, MilliporeSigma, Burlington, MA, USA), IgG antibody (trim away:0.5 mg/ml, #12-371, MilliporeSigma, Burlington, MA, USA). Goat anti-human-Alexa-633 (IF: 1:200; #A21091, Life Technologies, Carlsbad, CA, USA) and donkey anti-rabbit-Alexa-568 (IF: 1:200; #A10042, Life Technologies, Carlsbad, CA, USA), anti-alpha/beta-tubulin (IF: 1:100, #ATN02, Cytoskeleton, Denver, CO, USA), anti-HRB (WB: 1:1000; rabbit, #R1006, Kindle Biosciences, Greenwich, CT, USA) were used as secondary antibodies.

Immunofluorescence was performed as described previously (*67*). Briefly, the oocytes were fixed in 2% PFA at room temperature for 20 min. Blocking buffer (0.3% BSA in PBS with 0.01% Tween-20) was used to wash out the fixative solution three times for 10 min. Oocytes were then permeabilized in PBS containing 0.2% Triton-X-100 for 20 min and blocked in blocking buffer for 10 min. Primary antibodies incubation was performed for 1 h at room temperature in dark, humidified chamber, followed by three washes of 10 min each in blocking solution. Then oocytes were incubated in secondary antibody for 1 h in a dark humidified chamber, followed by three washes of 10 min each in blocking buffer. Finally, oocytes were mounted in 10 μl of VectaShield with DAPI.

### Oocyte microscopy and image analysis

Images were acquired using a Leica TCS SP8 confocal microscope equipped with a 63× objective, 1.40 N.A. oil immersion objective. For each image, optical z-slices were obtained using a 0.5-1 μm step length with a zoom setting of 2.5-4. Laser powers were kept constant for pixel intensity comparisons. Images were analyzed using NIH ImageJ software(*73*) or Imaris 9.9.1 from Bitplane (Zürich, Switzerland).

### AURKC localization analysis

Images of immunolabeled telophase oocytes were analyzed with Imaris 9.9.1. In Imaris, a line plot was constructed across the midzone spindle and the signal intensity representing AURKC was measured. Z-slices from each image were merged into a projection.

### Kinetochore-microtubule attachment assay

Oocytes matured for 8 h (Metaphase I) were transiently exposed to cold temperatures to depolymerize microtubules unattached to kinetochores. Oocytes were then immediately fixed in 2% PFA at room temperature for 20 min and blocked overnight. Oocytes were probed for alpha-tubulin and CREST and counterstained with DAPI. Oocytes were imaged using the protocol described above with the addition of Leica SP8 Lightning deconvolution. Images of immunolabeled cold-treated Metaphase I oocytes were analyzed in Imaris 10.1.1. Kinetochore foci were masked using the “spots” tool in Imaris. For each experiment, the median intensity of tubulin signal per kinetochore focus was used to determine whether each kinetochore overlapped with a microtubule bundle. Intensity thresholds were manually verified using at least 50+ kinetochores across 5+ oocytes per imaging set to account for inter-experimental variation.

### Statistical analysis

Unpaired Student’s t-test and one-way ANOVA based on the group numbers were used to evaluate the differences between groups using GraphPad Prism. The differences with p<0.05 were considered significant. The error bars indicate standard error of mean (S.E.M.). For the chromosome counting analysis of knock-in mice, oocyte data were pooled and analyzed as contingency outcomes via two-sided Fisher’s exact test.

Female fertility was modeled using nonlinear regression with an exponential plateau model. The similarity of the fertility between genotypes was evaluated using the extra sum-of-squares F Test. Multiple comparison corrections were performed using Tukey’s multiple comparison test. For both male and female trials, no outliers were identified as determined by <u>R</u>obust regression and <u>Out</u>lier removal (ROUT) analysis performed for each genotype at a Q coefficient of 1%.

## List of Supplementary Materials

### Seven supplementary figures

- Figure S1. Protein–protein interaction network of 107 candidate genes.
- Figure S2. Localization of KIF20A and KIF18A in Metaphase I Mouse Oocytes.
- Figure S3. Expression of exogenous GFP-tagged human kinesin variant proteins.
- Figure S4. AURKC signal at the midzone spindle in oocytes treated with KIF20A inhibitor.
- Figure S5. CRISPR strategy for generating the Kif18a-S273A mouse line.
- Figure S6. Kif18a variant male mouse fertility trial results.
- Figure S7. Consequences of exogenous expression of hKif18A variant in mitosis.

### Seven supplementary tables

- Table S1 Summary of individual phenotype, group assignment, and sequencing statistics
- Table S2 Top 404 genes (p<0.05) from the VAAST analysis
- Table S3 GO term enrichment of 404 candidate genes
- Table S4 PPI network interactions
- Table S5 GO terms from the modules identified from the PPI network
- Table S6 Candidate variant validation
- Table S7 CRISPR editing oligos and validation primers

### One supplementary data file

- Data S1. Variants identified in this study, along with their allele frequencies in the HRG and LRG groups.

## Data Availability

All data produced in the present study are available upon reasonable request to the authors

## Acknowledgments

The authors thank members of the Schindler and Xing labs for their helpful comments. This work was supported by NIH grant R01HD091331 to KS and JX. LB was supported by NIH fellowship F30HD107976, the Foundation for Women’s Wellness, and NIH grant T32GM139804. The Tolić lab is funded by the European Research Council (ERC Synergy Grant, GA Number 855158), the Croatian Science Foundation (HRZZ) through Swiss-Croatian Bilateral Projects (project IPCH-2022-10-9344), and projects co-financed by the Croatian Government and the European Union through the European Regional Development Fund—the Competitiveness and Cohesion Operational Programme: IPSted (Grant KK.01.1.1.04.0057) and QuantiXLie Center of Excellence (Grant KK.01.1.1.01.0004).

