## Supplementary figures and images for "Maternal genetic variants in kinesin motor domains prematurely increase egg aneuploidy"

### S1

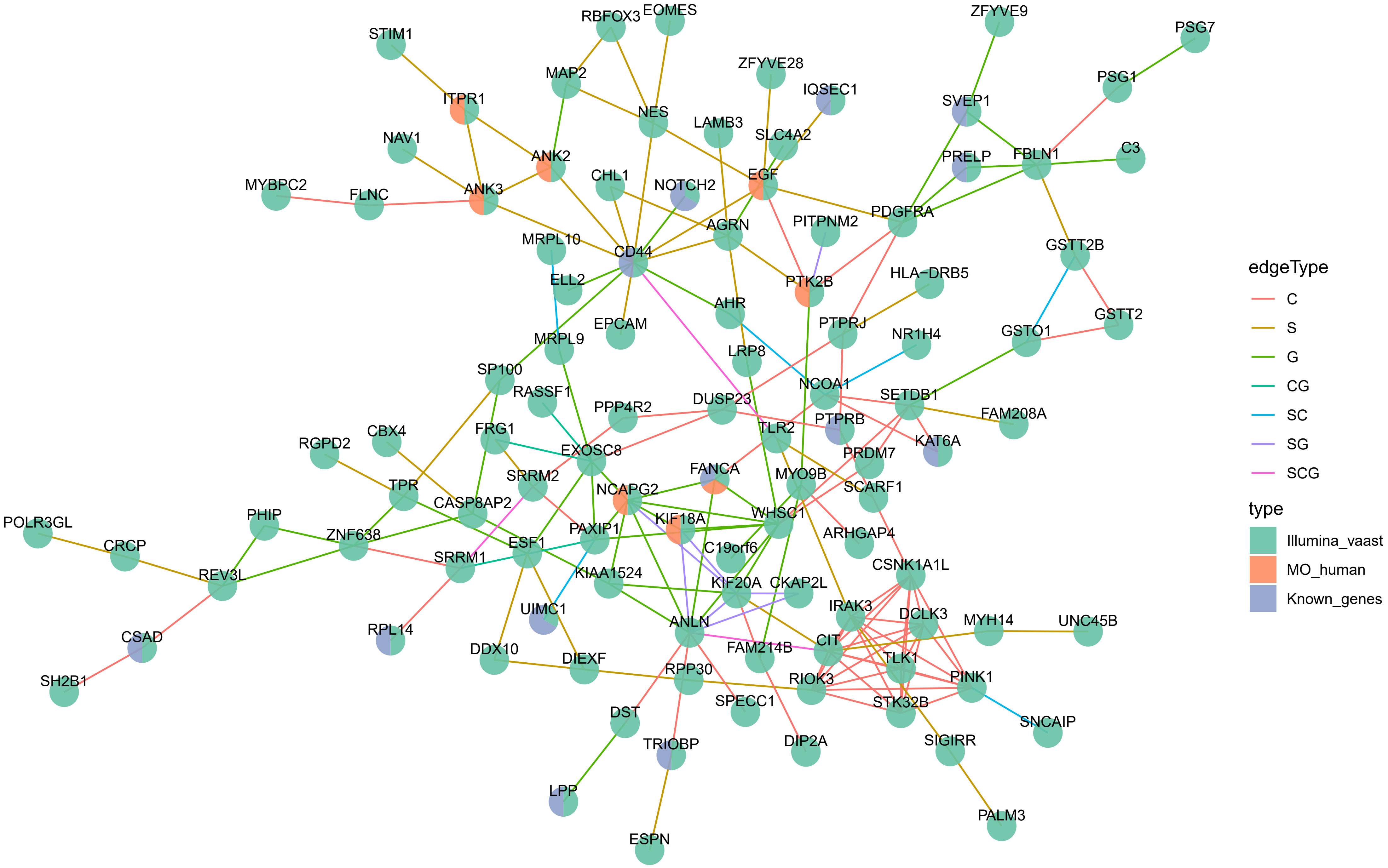

### S2

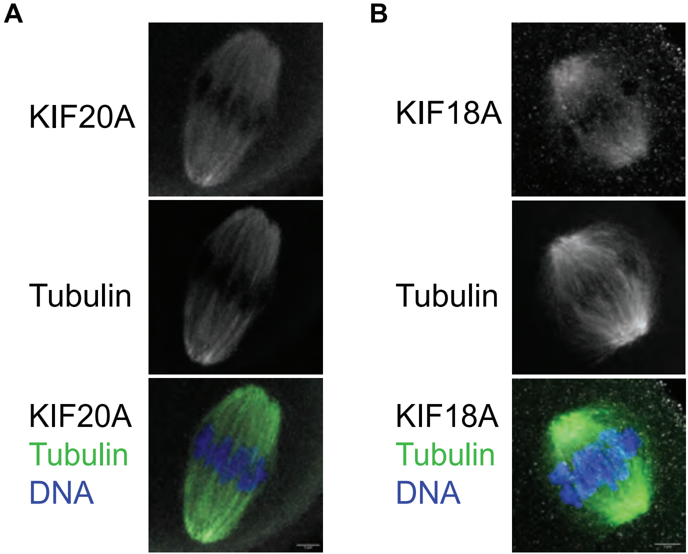

### S3

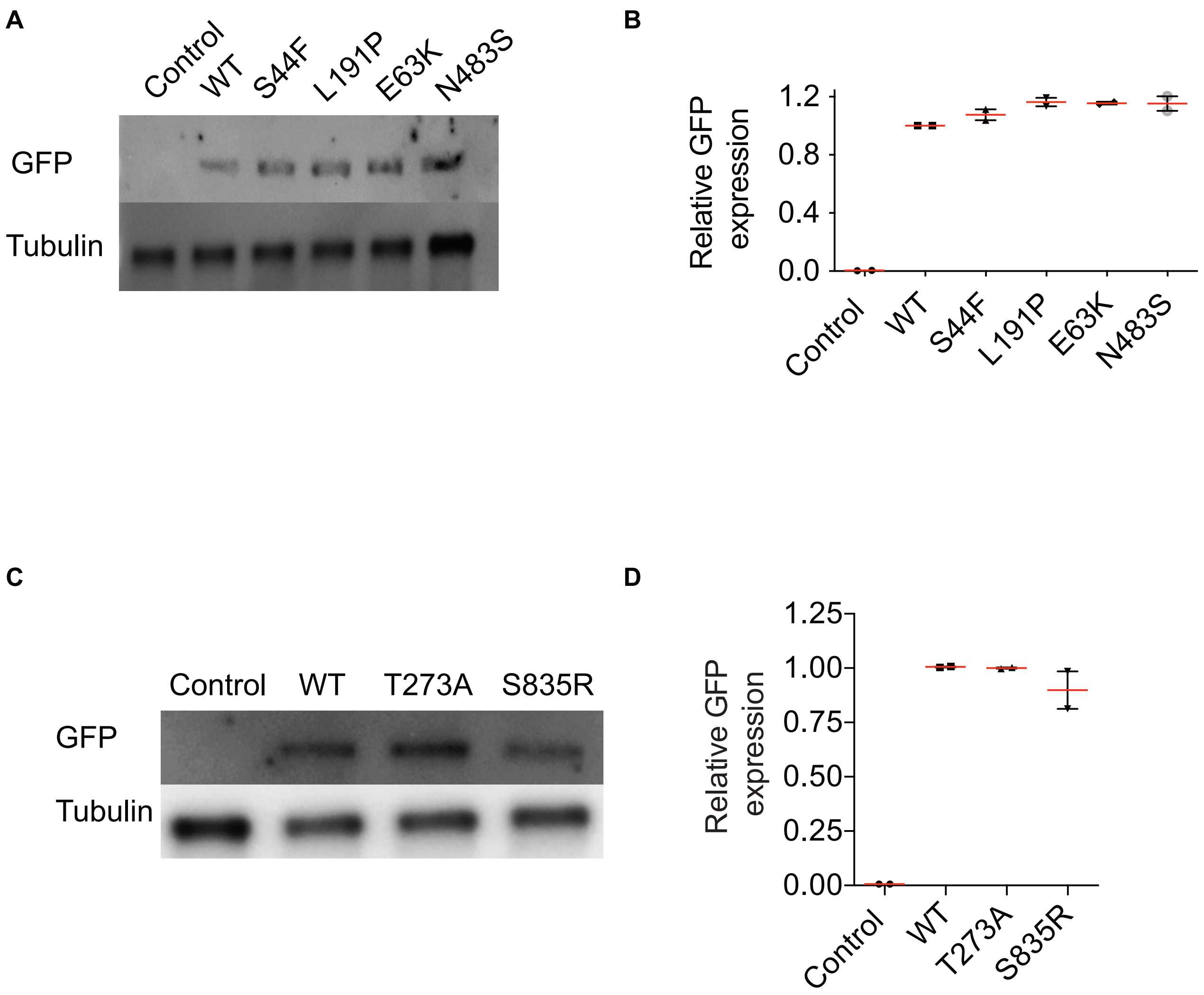

### S4

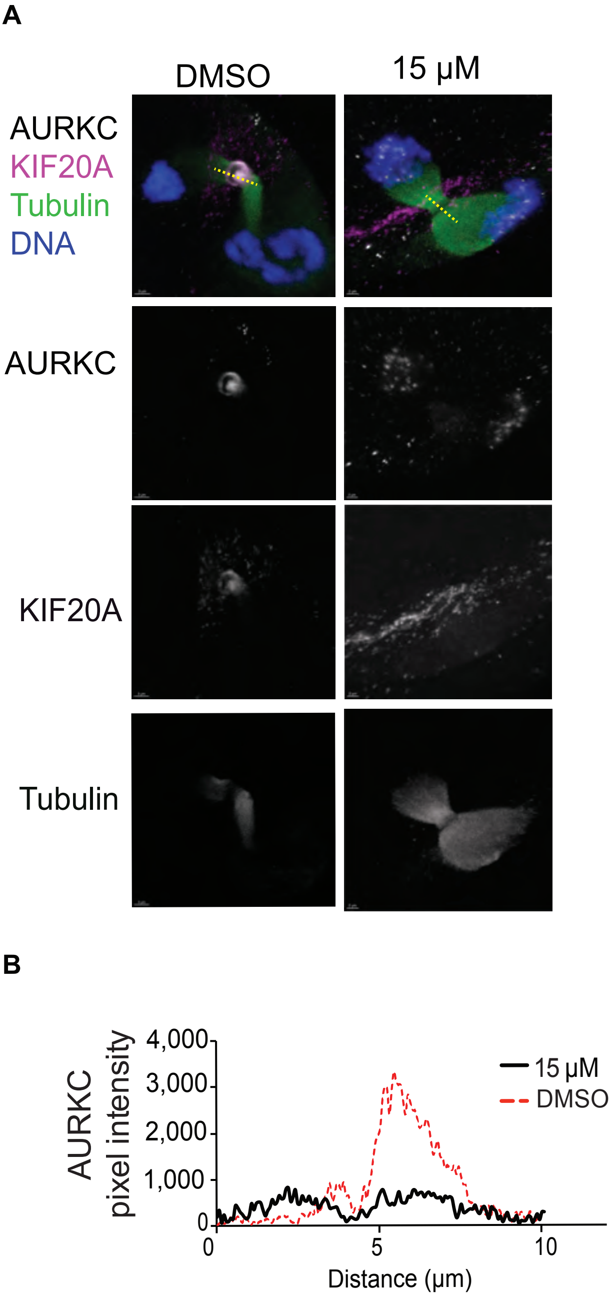

### S5

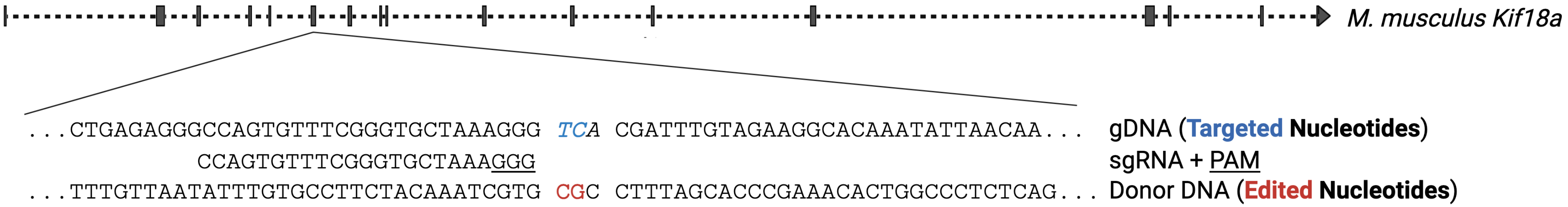

### S6

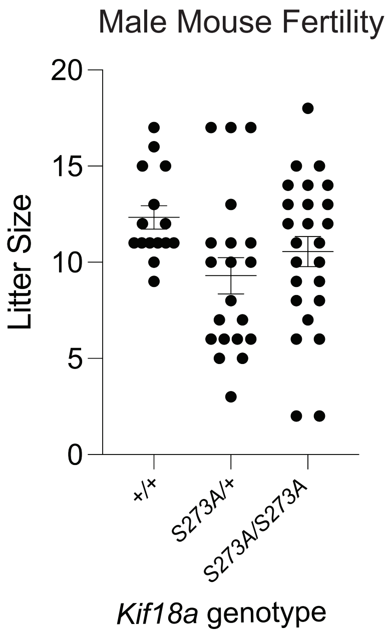

### S7

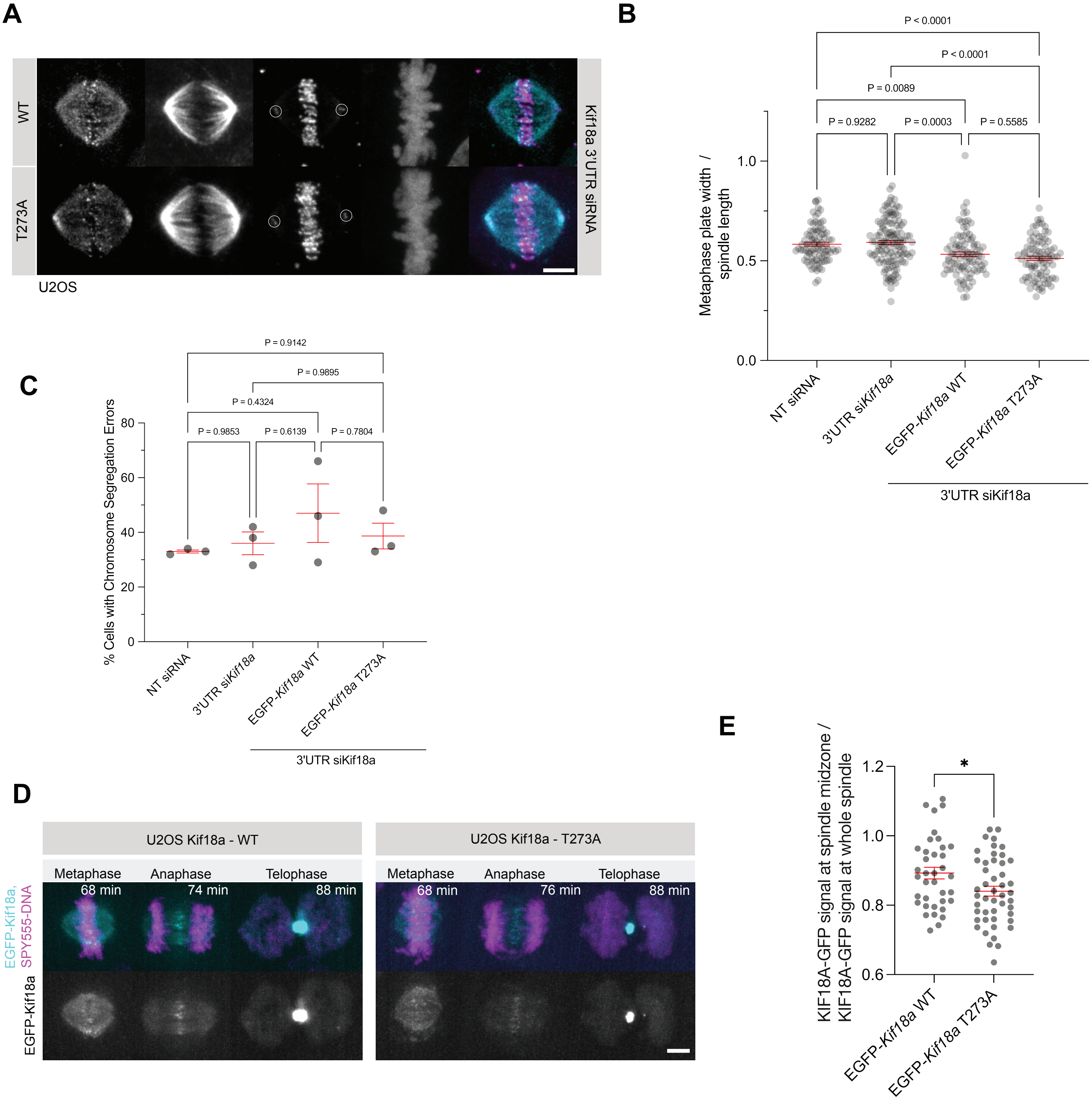
